# A retrospective analysis of the dynamic transmission routes of the COVID-19 in mainland China

**DOI:** 10.1101/2020.03.01.20029645

**Authors:** Xiandeng Jiang, Le Chang, Yanlin Shi

## Abstract

The fourth outbreak of the Coronaviruses, known as the COVID-19, has occurred in Wuhan city of Hubei province in China in December 2019. We propose a time-varying sparse vector autoregressive (VAR) model to retrospectively analyze and visualize the dynamic transmission routes of this outbreak in mainland China over January 31 - February 19, 2020. Our results demonstrate that the influential inter-location routes from Hubei have become unidentifiable since February 4, 2020, whereas the self-transmission in each provincial-level administrative region (location, hereafter) was accelerating over February 4-15, 2020. From February 16, 2020, all routes became less detectable, and no influential transmissions could be identified on February 18 and 19, 2020. Such evidence supports the effectiveness of government interventions, including the travel restrictions in Hubei. Implications of our results suggest that in addition to the origin of the outbreak, virus preventions are of crucial importance in locations with the largest migrant workers percentages (e.g., Jiangxi, Henan and Anhui) to controlling the spread of COVID-19.

## 1 Introduction

Coronaviruses are single-stranded, enveloped and positive-sense RNA viruses, which are spherical in shape and have petal-like spines [1]. Firstly discovered and identified in 1965 [2], coronaviruses have not caused large-scale outbreaks until the 2003 SARS epidemic in China, followed by 2012 MERS in Saudi Arabia and 2015 MERS in South Korea [3]. Although the exact origin remains debatable [4], the fourth outbreak has taken place in Hubei province of China in December 2019 and rapidly spread out nationally [5-10]. On January 10, 2020, the World Health Organization (WHO) temporarily named the new coronavirus as the 2019 novel coronavirus (2019-nCoV). Around one month later, the WHO officially renamed it to coronavirus disease 2019 or COVID-19 on 11 February, 2020 (see https://www.who.int/docs/default-source/coronaviruse/situation-reports/20200211-sitrep-22-ncov.pdf?sfvrsn=fb6d49b1_2 for details) and released a comprehensive interim guidance on dealing with this new virus for all countries [11]. On March 11, 2020, the WHO declared COVID-19 a global pandemic [12]. Since China reported its first cases to the WHO in December 2019, COVID-19 has been spreading rapidly around the world. As of June 15, 2020, about 7.9 million confirmed cases and 433 thousand deaths have been reported by authorities in 214 countries and territories [13].

To combat against the rapid spread of the COVID-19, since mid-January 2020, the central government of China and all local governments have implemented intensive preventions. Examples include tracing close contacts and quarantining infected cases, promoting social consensus on self-protection like wearing face mask in public area, among others [14]. With the unexpectedly rapidly growing number of confirmed cases, more extreme and unprecedented measures have taken places. On 23 January, 2020, the Chinese authorities introduced travel restrictions on five cities (Wuhan, Huanggang, Ezhou, Chibi and Zhi- jiang) of Hubei, shutting down the movement of more than 40 million people [15]. Among existing research, most argues that those interventions have effectively halted the spread of the COVID-19 [15-25].

The feasibilities of aforementioned stringent measures adopted by the Chinese government have been widely discussed, and existing studies have preliminarily examined the effectiveness of these local containment measures (e.g., the lockdown of Wuhan [26-28], airport screening and travel restrictions [19, 29-31], and isolation of cases and quarantine of contacts [32, 33]). Moreover, different types of infection control measures are enforced in many other countries to prevent and constrain the spread of the COVID-19, and the effects of these measures have been analyzed and compared over a rage of affected countries (e.g., Australia [34], Germany [35], Italy [36, 37] and South Korea [38]). Among those emerging large volume of studies, mathematical and statistical modeling plays a non-negligible role. Also, the classical susceptible exposed infectious recovered (SEIR) model with its various extensions is the most popular method [39-52]. SEIR family models are effective in exploring the epidemic characteristics of the outbreak, forecasting the inflection point and ending time, and deciding the measures to curb the spreading. Despite this, they are less appropriate in identifying transmission routes of the COVID-19 outbreak, which is also not thoroughly investigated in existing literature.

In this paper, we fill in this gap and perform a retrospective analysis using the publicly available data [53]. Rather than employing the SEIR, we develop a time-varying coefficient sparse vector autoregressive (VAR) model. Using the least absolute shrinkage and selection operator (lasso) [54, 55] and the local constant kernel smoothing estimator [56], our model is capable of estimating the dynamic high-dimensional Granger causality coefficient matrices. This enables the detection and visualization of time-varying inter-location and self-transmission routes of the COVID-19 on the daily basis. The resulting “road-map” can help policy-markers and public-health officers retrospectively evaluate both the effectiveness and unexpected outcomes of their interventions. Such an evaluation is critical to winning the current battle against COVID-19 in China, providing useful experience for other countries facing the emerging threat of this new coronavirus, and saving lives when a new epidemic occurs in the future.

## 2 Methods

### 2.1 Model

Throughout this study, we are interested in the growth rate *y_i_,_t_* such that:

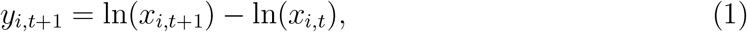

where *x_i_,_t_* is the accumulated confirmed cases in the provincial-level administrative region (location, hereafter) i on day *t (i* = 1,…,*N* and *t* = 1,…,*T*). *T* and *N* define the number of days and number of locations under consideration, respectively. We then define *y_t_* = (*y*_1_,*_t_*,…,*y_N_*,*_t_*}, an *N* × 1 vector of the growth rate on day t. To investigate a dynamic direct transmission of the growth rate among locations, we propose a time-varying coefficient sparse VAR model, namely the tvSVAR model, which assumes that Granger causality coefficients are functions of time, such that:

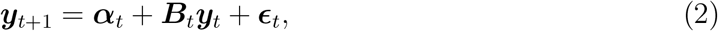

where ***α****_t_* is an *N*-dimensional intercept vector at time *t*. ***B****_t_* is an *N* × *N* Granger causality matrix at time *t* with a dynamic sparse structure, for which entries can be exactly zero and the locations of zeros can vary with time. ∊*_t_* is an *N* × 1 vector of error terms. The sparsity of ***B****_t_* is assumed because *N* could be even larger than T in our case, which leads to very unstable estimations and problematic interpretations of ***B****_t_*.

One important benefit of using the proposed tvSVAR to model the transmissions is that the Granger causality matrix, ***B****_t_*, can provide both the direction and strength of the route on day *t*. For example, the *ij^th^* entry in ***B****_t_* measures the strength of the transmission from location *i* to location j on day *t*. The *i^th^* diagonal of ***B****_t_* represents the self-transmission in location *i* that captures the relationship between the growth rate in the current and previous days. More critically, the sparse structure eases the interpretation of ***B****_t_* because many weak transmissions may be of a random nature. The corresponding coefficients, therefore, can be treated as noises and are shrunk to zeros exactly. Moreover, a time-varying design of ***B****_t_* allows us to investigate changes in the identified transmissions over time. For instance, let 11, 14 and 17 indicate Hubei, Jiangxi and Shanghai, respectively. On day *t* =1, the estimated *β*_11_,_14_,_1_ and *β*_14_,_17_,_1_ are 0.52 and 0.35, respectively. This suggests on that day, moderately strong transmission routes of confirmed COVID-19 cases are detected from Huber to Jiangxi and from Jiangxi to Shanghai, respectively. Further, estimated *β*_17,_*_i,_*_1_ for all *i* = 1,…, 20 are zeros, suggesting that the confirmed cases in Shanghai cannot spread to other locations on day 1. On day *t* = 2, we observe estimated *β*_11,14,1_ = 0.61, *β*_14,17,1_ = 0.41 and all *β*_17,_*_i,_*_1_ = 0. Thus, the two detected routes from Huber to Jiangxi and Jiangxi to Shanghai have become more influential, whereas the cases in Shanghai are still yet to spread out on day 2. The above results cannot be derived using the classic epidemiological SEIR model.

To capture both dynamic and sparse structure of the Granger causality coefficients, we solve the following optimization problem:

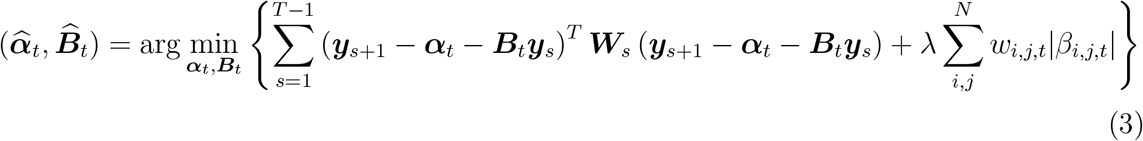

where 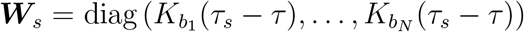 is the matrix of kernel weights calculated based on the bandwidth *b_i_, i* = 1,…, *N*, and 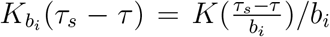 with *τ_s_* defined as a scaled time 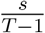. We use the Epanechnikov kernel *K*(*x*) = 0.75(1 − *x*^2^)_+_ and a unified bandwidth for each *i (b_i_* ≡ *b*) to avoid a large number of tuning parameters. The coefficients *β_i_,_j_,_t_* denotes the *ij^th^* entry of the Granger causality matrix ***B****_t_*, and *λ* is the tuning parameter that aims to shrink insignificant *β_i_,_j_,_t_* to zero and thus controls the sparsity of ***B****_t_*. Another essential feature of our proposed model is that the adaptive weights *w_i,j,t_* are employed to penalize *β_i,j,t_* differently in the lasso (L1) penalty [54, 55]. The choice of weights *w_i_,_j_,_t_* takes account of the prior knowledge about the transmissions and can be specified by the users. In this study, we consider *w_i,j,t_* as the reciprocal of the accumulated confirmed in location *i* on day *t −* 1. That is, the growth rate of a location with a smaller accumulated confirmed cases is less likely to influence the growth rates of others, and thus, more likely to be shrunk to zero. The final sparsity structure of ***B****_t_* is still data-driven.

The estimators as in (3) can also be viewed as a penalized version of local constant kernel smoothing estimator [56]. We utilize a modified version of the fast iterative soft thresholding algorithm (FISTA) [57] to solve the optimization problem (3).

Given a bandwidth *b* and a penalty parameter *λ*, we can find the estimator (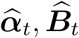) for each day t and observe the dynamic patterns of the transmission over time t for each pair of locations. The selection of b is critical to detecting the influential routes, which depends on the chosen criterion. Among the existing literature, a popular approach is to adopt the cross-validation strategy, such that based on the estimated (***α****_t_*, ***B****_t_)*, the model will not ‘overfit’ ***y****_t_*. As for the time-series analysis, we use an expanding-window sample to implement the cross-validation [58]. This requires that the chosen b will minimize the cross-validated forecast error, which is measured by the one-step-ahead root mean squared forecast error (RMSFE), such that

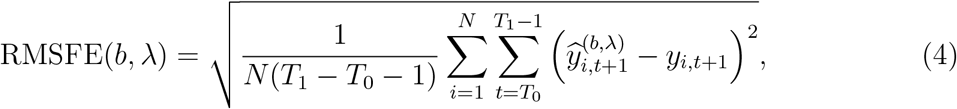

where [*T*_0_, *T*_1_] is the evaluation period, which is given by the last third of the data in our study, 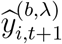 denotes the one-step-ahead forecast for location *i* based on the data up to day *t*, and *y_i,t_*_+1_ defines the observed growth rate at day *t* + 1 for location *i*. Note that RMSFE is analogous to the square root of the popular least squared errors. An interpretation is that the chosen b will lead to the minimized total out-of-sample forecast errors of the growth rates of confirmed cases over the last third of the sample period.

### 2.2 Code availability

The R code that supports the findings of this study is available from the author on request.

## 3 Data and Results

### 3.1 Data

The data studied in this paper include confirmed COVID-19 cases which occurred in mainland China. The data are publicly available and sourced from the website of the National Health Commission of the People’s Republic of China [53]. The data-coverage ranges from January 29, 2020 to February 19, 2020, during which no missing data were recorded at location-level. The accumulated cases and the associated growth rates, grouped by the total national number, cases in Hubei and cases in all other locations, are plotted in Figure 1 (a) and (b), respectively. The total national (Hubei) accumulated confirmed cases increased rapidly from 7,736 (4,586) on January 31, 2020 to 75,101 (62,457) on February 19, 2020. Note that on February 12, 2020, confirmed cases in Hubei included those confirmed by both laboratory and clinical diagnosis, leading to a one-time hump of the accumulated number. Compared to those of Hubei, confirmed cases of other locations took up a smaller proportion of the total national number, ranging from 40.7% on January 29, 2020 to 16.8% on February 19, 2020. This suggests that the growth rate of other locations should be lower than that of Hubei, which is consistent with Figure 1 (b). Throughout our investigation period, except for the one-time hump on February 12, 2020, growth rates of Hubei and the rest steadily declined, from 33% and 25% to 5% and 1%, respectively.

**Figure 1:**
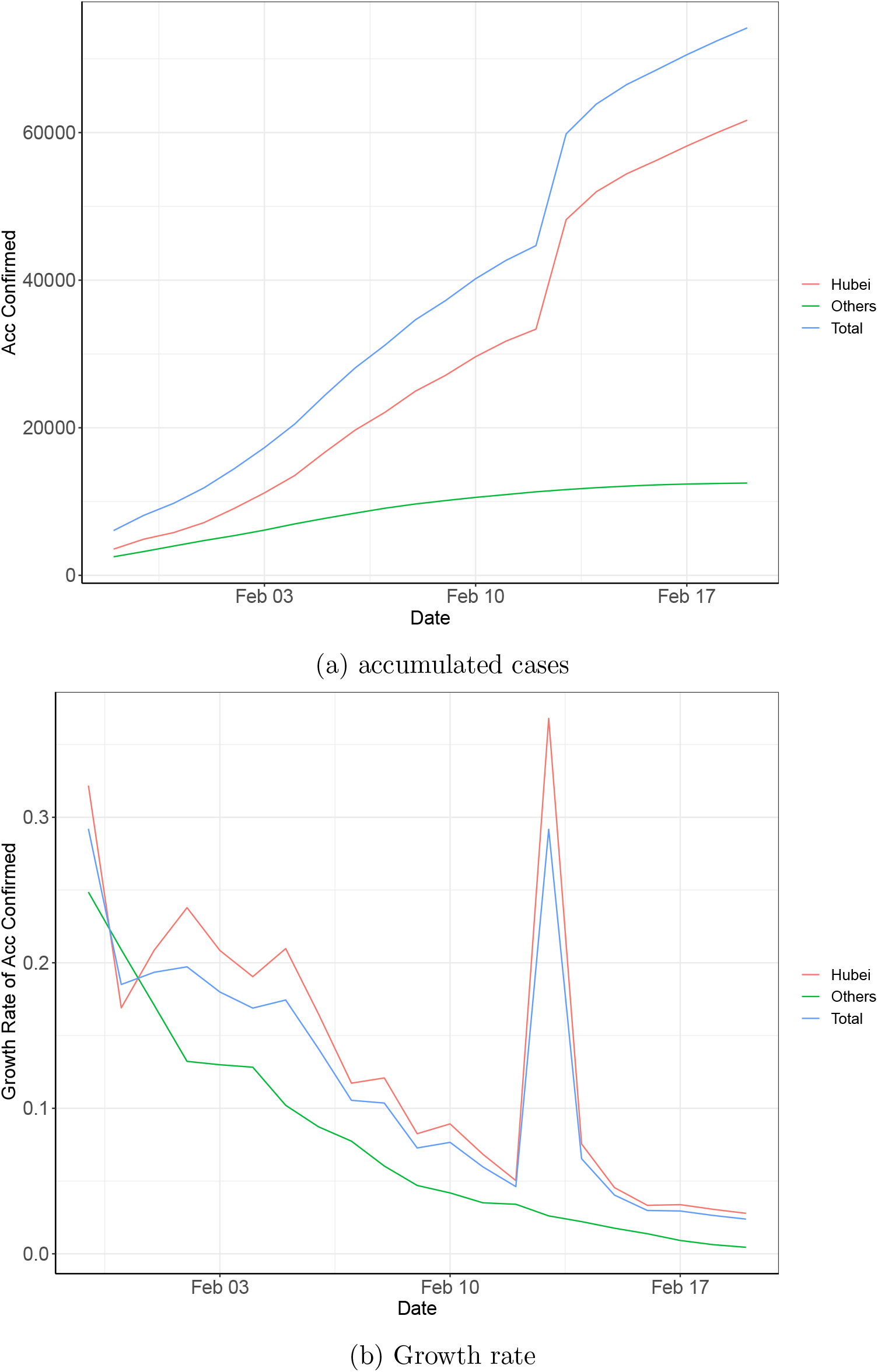
Accumulated confirmed cases and growth rate: 31/1/2020-19/2/2020

### 3.2 Estimation results: Transmission routes

By taking the difference of the logged accumulated cases and applying one lag, our estimated transmission routes are available from January 31 to February 19, 2020 (two observations are lost). To avoid potential noises caused by small numbers, we only include data of locations, which had at least 150 accumulated confirmed cases as of February 19, 2020. Altogether, our modeled sample contains 20 location-level confirmed cases. We firstly test the stationarity of the 20 growth rates separately. Based on the Augmented Dickey-Fuller test, only the rates of 3 locations (Beijing, Hainan and Heilongjiang) are insignificant, which is in-line with the employed 10% significance level. The detailed results are available upon request. The model explained in Section 2 is then fitted incorporating all the 20 growth rates. A non-zero estimate of *β_i,j,t_*, the *ij^th^* entry of ***B****_t_* in (2), indicates that on the *t^th^* day, the growth rate of location *j* is Granger caused by that of location *i*. In other words, there is a transmission route from location *i* to location *j*. Among the 20-day results, we noticed that the estimated transmission routes on days 1-5 changed considerably on daily basis. From the sixth day onwards, however, those estimated routes were more steady.

Hence, we plot the estimates on days 1-5 and those on the every fifth day thereafter, on Figure 2. Be noted that estimates smaller than 0.2 (none-influential) are not presented a better visual illustration purpose. Also, this research focuses on the analysis of mainland China only, which excludes Taiwan, Macau, Hong Kong and all important islands of China’s territory, such as those located in the South China Sea. The plots presented in Figure 2 therefore do not present a complete map of the territory of China, nor should they be used for purposes other than displaying identified transmission routes of COVID-19 in mainland China. The readers are directed to The National Administration of Surveying, Mapping and Geographic Information of the People’s Republic of China, should they need to precisely explore the scope of maps, national boundaries and the drawing of important islands of the Chinese territory.

**Figure 2:**
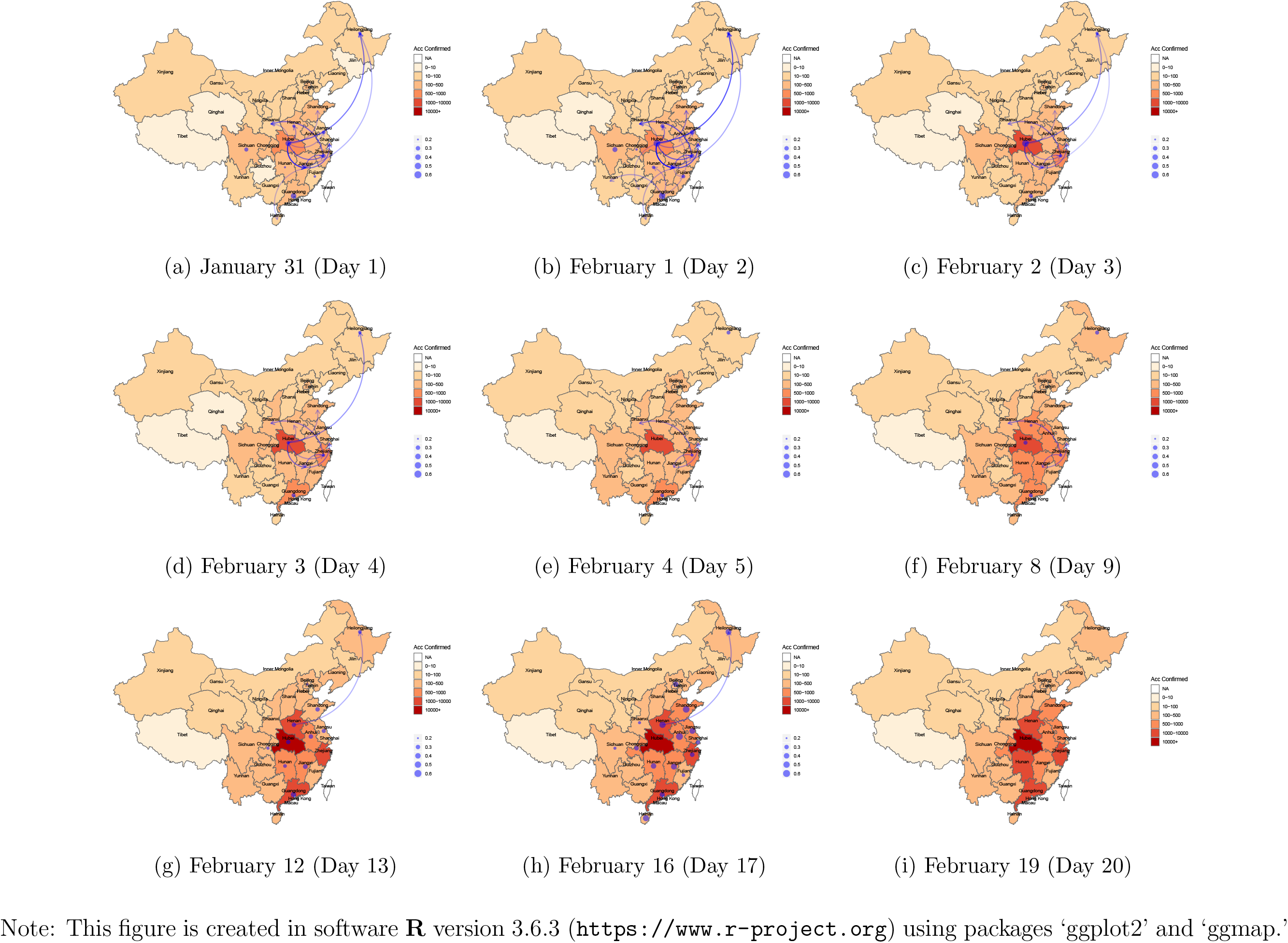
Estimated routes of transmission among locations of China: 31/1/2020-19/2/2020.

In Figure 2, we use color of light orange (small) to dark red (large) indicating the accumulated confirmed cases in each location, up to time t. Estimated transmission routes are colored in blue. Self-transmissions (indicated by *β_i_,_i,t_*) are denoted by dots, and a larger size of dot suggests a larger estimated *β_i_,_i_,_t_*. Inter-location transmission (indicated by *β_i,j,t_*, where *i* ≠ *j*) is represented by arrows, with the transparency indicating the magnitude of estimated *β_i,j,t_*. On the first day (January 31, 2020), there were influential inter-location transmissions from Hubei to Jiangxi, Heilongjiang, Zhejiang, Henan, Shandong, Jiangsu and Shaanxi, sorted by the magnitudes of strength (big to small). There were a few additional detected such transmissions on the second day, including those from Hubei to Guangxi, from Jiangxi to Fujian, and from Guangdong to Anhui, Yunnan and Hunan. The number of such identified inter-location routes, however, reduced rapidly over the next three days. On the fifth day (February 4, 2020), no influential transmission routes were found from Hubei to directly affect other locations, and there were only three influential routes identified nationally, including Zhejiang–Shaanxi, Zhejiang–Jiangxi and Jiangxi–Shanghai. The number of those detected inter-location routes declined again in the next few days, and on day 13, only Henan–Heilongjiang was found influential. On days 19 and 20 (February 18 and 19, 2020), there were no influential inter-location transmissions identified. The above findings suggest that the number of influential inter-location transmissions overall dropped quickly in the first five days and then reduced steadily for the rest fifteen days. This is consistent with the observations of Figure 3 (a), where the time-varying estimates of the Granger causality of Hubei on other locations are plotted. On each day, we report the mean, standard deviation (Std. Dev.), the 25% quantile (*Q*_1_) and 75% quantile (*Q*_3_) of those estimates in Table 1, which also leads to consistent findings.

**Figure 3:**
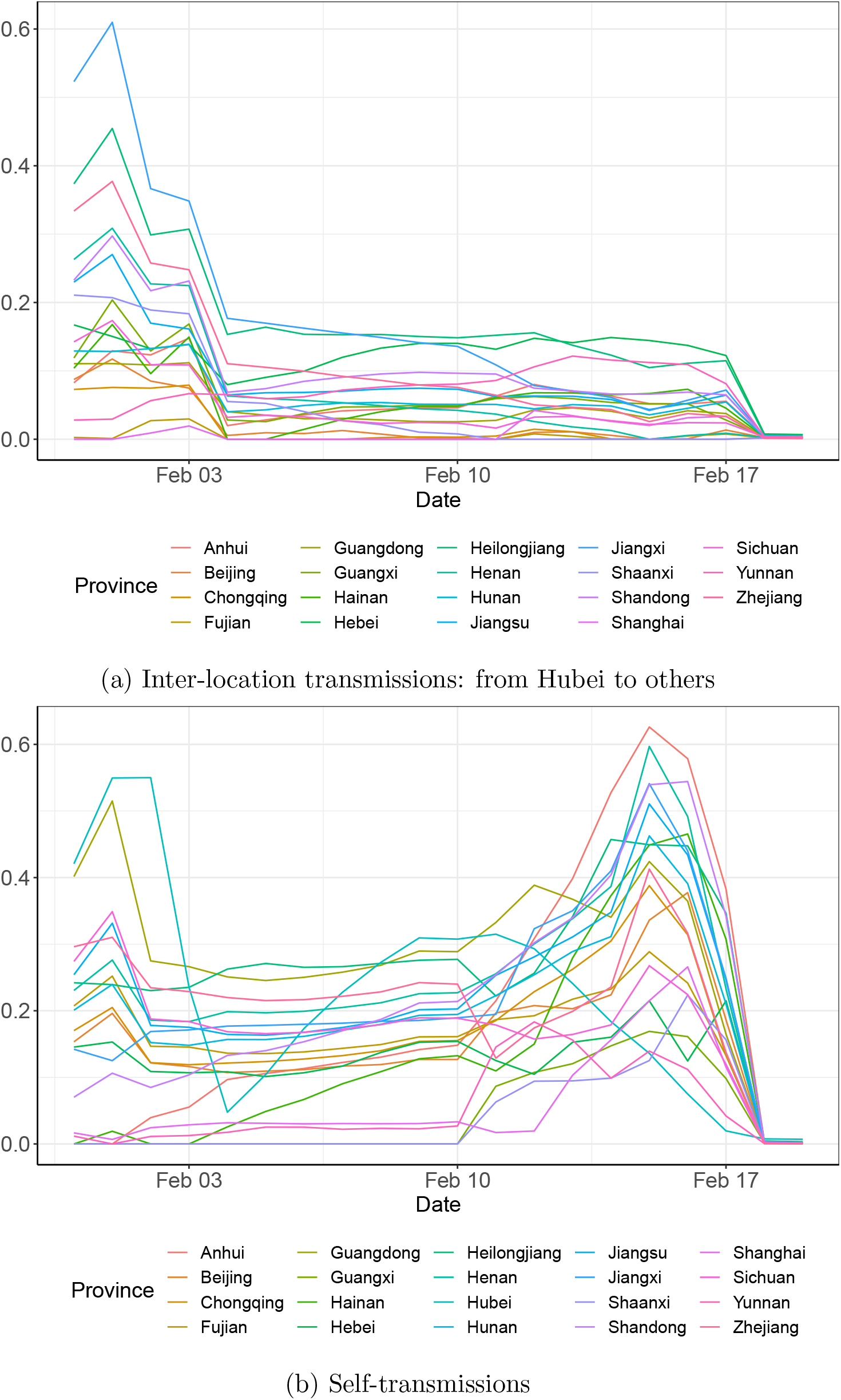
Estimated time-varying coefficients: 31/1/2020{19/2/2020

**Table 1:**
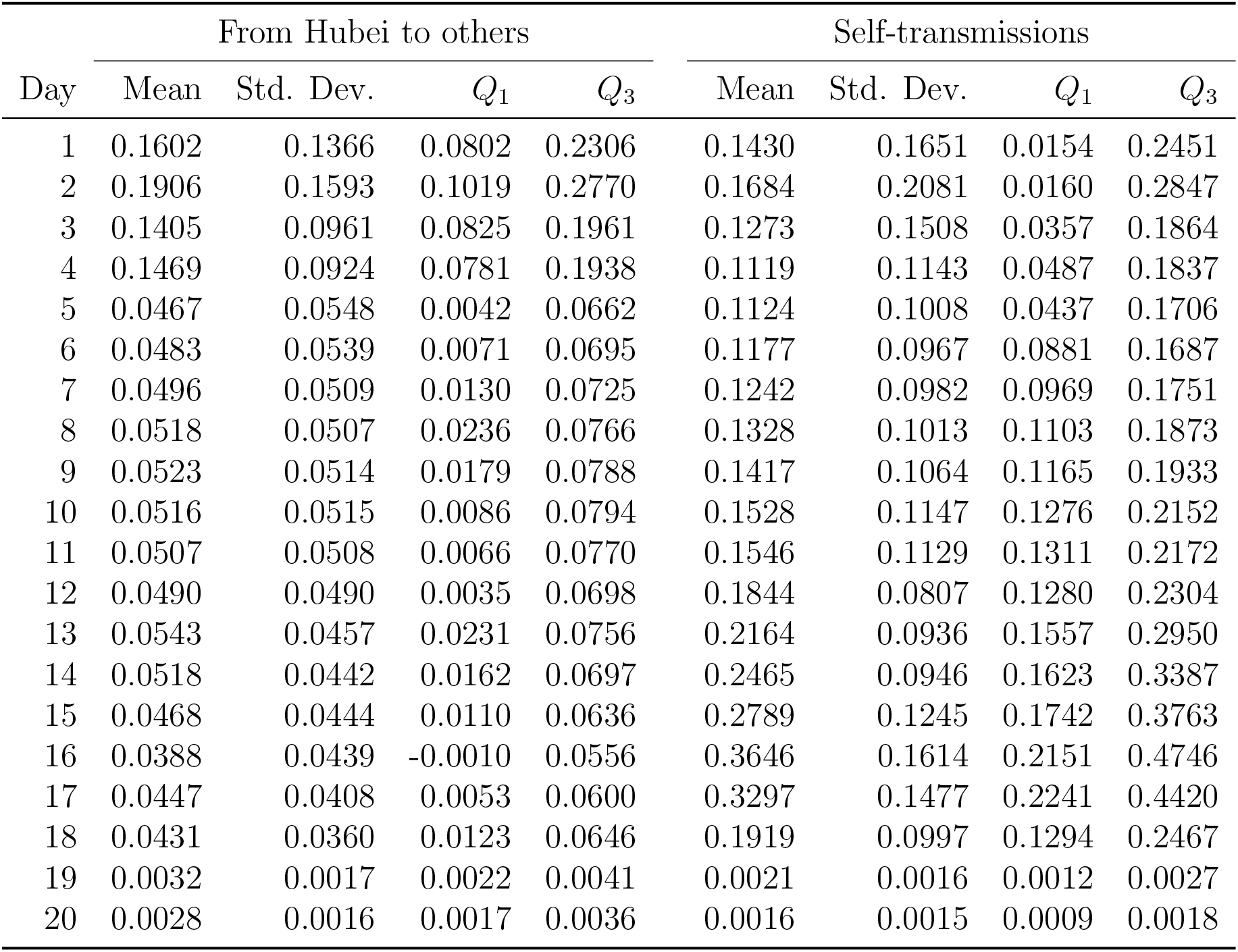
Summary of daily estimated transmission routes

As for the self-transmission, we firstly examine Figure 2 (b). It can bee seen that there were quite a few detected influential self-transmissions on the first two days. However, this number dropped quickly over days 3–5, and only self-transmissions of Heilongjiang, Guangdong and Zhejiang were found influential on day 5. Since then, the number of influential self-transmissions increased quickly with growing magnitudes (influence). On the sixteenth day (February 15, 2020), 16 out of the 20 examined locations had an estimated *β_i,i,_*_20_ of at least 0.2. Those large self-transmissions, however, disappeared rapidly again in the next three days. On February 18 and 19, 2020, there were no influential self-transmissions identified. This is consistent with our findings on Figure 3 (b), where time-varying estimated *β_i,i,t_* are plotted for each location. We report daily descriptive statistics of those estimates in Table 1, which also results in consistent conclusions.

## 4 Discussions

Since 23 January, 2020, many cities on mainland China started to introduce travel restrictions, including five cities (Wuhan, Huanggang, Ezhou, Chibi and Zhijiang) of Hubei [15]. According to WHO’s situation report [59], the average incubation period of COVID-19 is up to 10 days. Thus, our estimated dynamic transmission routes supports the significant effectiveness of the interventions taken by the Chinese authorities [15–25]. This is evidenced by Figure 2 (a)–(e), where the number of influential inter-location transmissions from Hubei to other locations reduced very quickly. Compared to multiple influential routes originating in Hubei detected on the first two days (January 31 and February 1, 2020), by February 4, 2020 (around 10 days after the travel restrictions), there were already no such transmissions identified. On the other hand, from February 5 to 16, 2020, Table 1 suggests that the averaged magnitudes of self-transmission on each day were strengthening steadily. This may also be explained by the interventions, which have effectively blocked inter-location transmissions, such that the growth rate of each location could only be caused by its internal transmissions.

We now focus on the inter-location transmission routes. Since influential routes from Hubei were no longer detected since day 5, we calculate the average *β_i,j,t_* of the 19 locations affected by Hubei over the first four days (January 31 – February 3, 2020). The top five destinations are presented in Panel A of Table 2. Apart from its geographic neighbors Jiangxi and Henan, Hubei has lead to influential routes to Heilongjiang, Zhejiang and Shandong directly. This may be explained by the substantial floating population working and living in Hubei from those locations. Excluding routes originating in Hubei, the Panel B of Table 2 suggests that the top destinations of transmission routes have not changed much over the first four days and the rest sixteen days. Despite minor differences in ranking, Shaanxi, Heilongjiang, Jiangxi, Anhui, Henan and Jiangsu appear to be the destinations suffered most from inter-location transmissions from origins other than Hubei. Similarly, the top five sources (excluding Hubei) of those transmissions are basically identical over the days 1–4 and days 5–20, as shown in Panel C of Table 2. This is consistent with the fact that travel restrictions in Hubei should not affect the connections among other locations. In all cases, Jiangxi, Henan, Guangdong, Zhejiang and Anhui are the most influential origins other than Hubei.

**Table 2:** Top five locations of the inter-location transmissions

| Days | High – Low |  |  |  |  |
| --- | --- | --- | --- | --- | --- |
| *Panel A: top destinations (affected by Hubei)* |  |  |  |  |  |
| 1–4 | Jiangxi | Heilongjiang | Zhejiang | Henan | Shandong |
| *Panel B: top destinations (affected by locations excluding Hubei)* |  |  |  |  |  |
| All | Shaanxi | Heilongjiang | Jiangxi | Anhui | Henan |
| 1-4 | Shaanxi | Jiangxi | Heilongjiang | Henan | Jiangsu |
| 5-20 | Heilongjiang | Shaanxi | Jiangxi | Anhui | Henan |
| *Panel C: top origins (excluding Hubei)* |  |  |  |  |  |
| All | Jiangxi | Henan | Guangdong | Zhejiang | Anhui |
| 1-4 | Jiangxi | Guangdong | Zhejiang | Henan | Anhui |
| 5-20 | Henan | Jiangxi | Guangdong | Anhui | Zhejiang |

It is worth noting that Jiangxi, Henan and Anhui belong to both the top origins and destinations of the inter-location transmissions, excluding Hubei. Since the impact of Hubei is not considered, this cannot be explained by the two influential transmission routes of Hubei–Jiangxi and Hubei–Henan listed in Panel A of Table 2. To see this, over days 5–20, the transmissions out of Hubei are no longer significant and thus should not affect routes from Jiangxi and Henan to another location. In contrast, one explanation is the large migrant workers from Jiangxi, Henan and Anhui to other locations (excluding Hubei). According to the Report on China’s migrant population development of 2017 [60], Jiangxi (7.25%), Henan (6.30%) and Anhui (6.27%) are among the top five locations in mainland China, ranked by the percentages of migrant workers in 2017.

## 5 Conclusions

Coronaviruses have lead to three major outbreaks ever since the SARS occurred in 2003. Although the exact origin is still debatable, the current shock, namely COVID-19, has taken place in Wuhan, the capital city of Hubei province in mainland China. As the fourth large-scale outbreak of coronaviruses, COVID-19 is spreading quickly to all provincial-level administrative regions (locations, hereafter) in China and has recently become a world-wide epidemic. As a significant complement to existing research, this study employs a tvSVAR model and retrospectively investigates and visualizes the transmission routes in mainland China.

Demonstrated in Figure 2, our baseline results review both the dynamic inter-location and self-transmission routes. Since February 4, 2020, the spread out of Hubei was largely reduced, leading to no identifiable routes to other locations. Simultaneously, the self-transmissions started to accelerate and peaked on around February 15, 2020 for most locations. Given an average incubation period of 10 days, those results support the argued effectiveness of the travel restrictions to control the spread of COVID-19, which took place in multiple cities of Hubei on January 23, 2020. On February 18–9, 2020, there existed no influential inter-location or self-transmission routes. Thus, the growth rates of confirmed cases are of a more random nature in all locations thereafter, implying that the spread of COVID-19 has been under control. For the detected inter-location transmissions, our findings demonstrate that Jiangxi, Heilongjiang, Zhejiang, Henan and Shandong are the top 5 locations affected mostly via routes directly from Hubei. When the influence of Hubei is excluded, Jiangxi, Henan and Anhui are among both the top origins and destinations of transmission routes.

Our results have major practical implications for public health decision- and policymakers. For one thing, the implemented timely ad-hoc public health interventions are proven effective, including contact tracing, quarantine and travel restrictions. For another, apart from the origin of the virus, as locations with largest migrant workers percentages, virus preventions are also of crucial importance in Jiangxi, Henan and Anhui to controlling the epidemics like the outbreak of COVID-19 in the future. With limited resources, taking ad-hoc interventions in such locations may most effectively help stop the spread of a new virus, from an economic perspective.

## Data Availability

Data are sourced from the R package R2019nCoV.

## Acknowledgment

The authors would like to thank the Southwestern University of Finance and Economics, Australian National University and Macquarie University for their support. Xiandeng Jiang acknowledges the research grants supported by the People Republic of China Ministry of Education Youth Project for Humanities and Social Science Research (grant No. 20YJC790051). We particularly thank the Deputy Editor (Rafal Marszalek) and two anonymous referees for providing valuable and insightful comments on earlier drafts. The usual disclaimer applies.

## Author Contributions

X.J. collected data and designed the research. L.C. and Y.S. performed the research and analyzed the data. X.J., L.C., and Y.S. wrote the paper.

## References

1. Chen, Y., Liu, Q. & Guo, D. Emerging coronaviruses: genome structure, replication, and pathogenesis. Journal of medical virology 92, 418–423 (2020).

2. Kahn, J. S. & McIntosh, K. History and recent advances in coronavirus discovery. The Pediatric Infectious Disease Journal 24, S223–S227 (2005).

3. Bogoch, I. I. et al. Pneumonia of unknown aetiology in Wuhan, China: potential for international spread via commercial air travel. Journal of travel medicine 27, taaa008 (2020).

4. Zhou, P. et al. A pneumonia outbreak associated with a new coronavirus of probable bat origin. Nature, 1–4 (2020).

5. Wu, F. et al. A new coronavirus associated with human respiratory disease in China. Nature, 1–5 (2020).

6. Huang, C. et al. Clinical features of patients infected with 2019 novel coronavirus in Wuhan, China. The Lancet 395, 497–506 (2020).

7. Cohen, J. & Normile, D. New SARS-like virus in China triggers alarm. Science 367, 234–235 (2020).

8. Lu, H., Stratton, C. W. & Tang, Y.-W. Outbreak of Pneumonia of Unknown Etiology in Wuhan China: the Mystery and the Miracle. Journal of Medical Virology 92, 401–402 (2020).

9. Parry, J. China coronavirus: cases surge as official admits human to human transmission. BMJ 368. eprint: https://www.bmj.com/content/368/bmj.m236.full.pdf. https://www.bmj.com/content/368/bmj.m236 (2020).

10. Li, G. & De Clercq, E. Therapeutic options for the 2019 novel coronavirus (2019-nCoV) 2020.

11. World Health Organization (WHO). https://www.who.int/healthtopics/coronavirus. 2020. (2020).

12. World Health Organization (WHO). https://www.euro.who.int/en/health-topics/health-emergencies/coronavirus-covid-19/news/news/2020/3/who-announces-covid-19-outbreak-a-pandemic. 2020. (2020).

13. Johns Hopkins University Center for Systems Science and Engineering. https://coronavirus.jhu.edu/map.html). 2020. (2020).

14. Peng, L., Yang, W., Zhang, D., Zhuge, C. & Hong, L. Epidemic analysis of COVID-19 in China by dynamical modeling. *arXiv preprint arXiv:2002.06563* (2020).

15. Tang, B. et al. Estimation of the Transmission Risk of the 2019-nCoV and Its Implication for Public Health Interventions. Journal of Clinical Medicine 9, 462 (2020).

16. Tang, B. et al. An updated estimation of the risk of transmission of the novel coronavirus (2019-nCov). Infectious disease modelling 5, 248–255 (2020).

17. Shen, M., Peng, Z., Guo, Y., Xiao, Y. & Zhang, L. Lockdown may partially halt the spread of 2019 novel coronavirus in Hubei province, China. *medRxiv* (2020).

18. Li, X., Zhao, X. & Sun, Y. The lockdown of Hubei Province causing different transmission dynamics of the novel coronavirus (2019-nCoV) in Wuhan and Beijing. *medRxiv* (2020).

19. Chinazzi, M. et al. The effect of travel restrictions on the spread of the 2019 novel coronavirus (COVID-19) outbreak. Science 368, 395–400 (2020).

20. Backer, J. A., Klinkenberg, D. & Wallinga, J. Incubation period of 2019 novel coronavirus (2019-nCoV) infections among travellers from Wuhan, China, 20-28 January 2020. Eurosurveillance 25 (2020).

21. Lai, S. et al. Assessing spread risk of Wuhan novel coronavirus within and beyond China, January-April 2020: a travel network-based modelling study. *medRxiv* (2020).

22. Quilty, B. J., Clifford, S., et al. Effectiveness of airport screening at detecting travellers infected with novel coronavirus (2019-nCoV). Eurosurveillance 25 (2020).

23. Clifford, S. J. et al. Interventions targeting air travellers early in the pandemic may delay local outbreaks of SARS-CoV-2. *medRxiv* (2020).

24. Hellewell, J. et al. Feasibility of controlling 2019-nCoV outbreaks by isolation of cases and contacts. *medRxiv* (2020).

25. Jin, G., Yu, J., Han, L. & Duan, S. The impact of traffic isolation in Wuhan on the spread of 2019-nCov. *medRxiv* (2020).

26. Yuan, C. et al. A simple model to assess Wuhan lock-down effect and region efforts during COVID-19 epidemic in China Mainland. *medRxiv* (2020).

27. Li, X., Zhao, X. & Sun, Y. The lockdown of Hubei Province causing different transmission dynamics of the novel coronavirus (2019-nCoV) in Wuhan and Beijing 2020.

28. Tian, H. et al. An investigation of transmission control measures during the first 50 days of the COVID-19 epidemic in China. Science 368, 638–642 (2020).

29. Anderson, R. M., Heesterbeek, H., Klinkenberg, D. & Hollingsworth, T. D. How will country-based mitigation measures influence the course of the COVID-19 epidemic? The Lancet 395, 931–934 (2020).

30. Kraemer, M. U. et al. The effect of human mobility and control measures on the COVID-19 epidemic in China. Science 368, 493–497 (2020).

31. Wells, C. R. et al. Impact of international travel and border control measures on the global spread of the novel 2019 coronavirus outbreak. Proceedings of the National Academy of Sciences 117, 7504–7509 (2020).

32. Hellewell, J. et al. Feasibility of controlling COVID-19 outbreaks by isolation of cases and contacts. The Lancet Global Health 8, e488–e496 (2020).

33. The effectiveness of quarantine and isolation determine the trend of the COVID-19 epidemics in the final phase of the current outbreak in China. International Journal of Infectious Diseases 95, 288–293 (2020).

34. Chang, S. L., Harding, N., Zachreson, C., Cliff, O. M. & Prokopenko, M. Modelling transmission and control of the COVID-19 pandemic in Australia. *arXiv preprint arXiv:2003.10218* (2020).

35. Dehning, J. et al. Inferring change points in the spread of COVID-19 reveals the effectiveness of interventions. Science 369 (2020).

36. Gatto, M. et al. Spread and dynamics of the COVID-19 epidemic in Italy: Effects of emergency containment measures. Proceedings of the National Academy of Sciences 117, 10484–10491 (2020).

37. Giordano, G. et al. Modelling the COVID-19 epidemic and implementation of population-wide interventions in Italy. Nature Medicine, 1–6 (2020).

38. Shim, E., Tariq, A., Choi, W., Lee, Y. & Chowell, G. Transmission potential and severity of COVID-19 in South Korea. International Journal of Infectious Diseases 93, 339–344 (2020).

39. Chen, T. et al. A mathematical model for simulating the transmission of Wuhan novel Coronavirus. *bioRxiv* (2020).

40. Chen, Y., Cheng, J., Jiang, Y. & Liu, K. A time delay dynamical model for outbreak of 2019-nCoV and the parameter identification. *arXiv preprint arXiv:2002.004 1 8* (2020).

41. Huang, N. E. & Qiao, F. A data driven time-dependent transmission rate for tracking an epidemic: a case study of 2019-nCoV. Science Bulletin 65, 425 (2020).

42. Jung, S.-m. et al. Epidemiological identification of a novel infectious disease in real time: Analysis of the atypical pneumonia outbreak in Wuhan, China, 2019-20. *medRxiv* (2020).

43. Lin, Q., Hu, T. & Zhou, X.-H. Estimating the daily trend in the size of COVID-19 infected population in Wuhan. *medRxiv* (2020).

44. Nishiura, H. et al. Estimation of the asymptomatic ratio of novel coronavirus infections (COVID-19). *medRxiv* (2020).

45. Nishiura, H et al. The Extent of Transmission of Novel Coronavirus in Wuhan, China, 2020. Journal of clinical medicine 9 (2020).

46. Read, J. M., Bridgen, J. R., Cummings, D. A., Ho, A. & Jewell, C. P. Novel coronavirus 2019-nCoV: early estimation of epidemiological parameters and epidemic predictions. *medRxiv* (2020).

47. Sanche, S. et al. The Novel Coronavirus, 2019-nCoV, is Highly Contagious and More Infectious Than Initially Estimated. *arXiv preprint arXiv:2002.03268* (2020).

48. Wu, J. T., Leung, K. & Leung, G. M. Nowcasting and forecasting the potential domestic and international spread of the 2019-nCoV outbreak originating in Wuhan, China: a modelling study. The Lancet 395, 689–697 (2020).

49. Xiong, H. & Yan, H. Simulating the infected population and spread trend of 2019-nCov under different policy by EIR model. *Available at SSRN 3537083* (2020).

50. Yang, Y. et al. Epidemiological and clinical features of the 2019 novel coronavirus outbreak in China. *medRxiv* (2020).

51. Zeng, T., Zhang, Y., Li, Z., Liu, X. & Qiu, B. Predictions of 2019-nCoV Transmission Ending via Comprehensive Methods. *arXiv preprint arXiv:2002.04945* (2020).

52. Zhao, S. et al. Preliminary estimation of the basic reproduction number of novel coronavirus (2019-nCoV) in China, from 2019 to 2020: A data-driven analysis in the early phase of the outbreak. International journal of infectious diseases 92, 214–217 (2020).

53. National Health Commission of the People’s Republic of China. http://www.nhc.gov.cn/xcs/xxgzbd/gzbd_index.shtml. 2020. (2020).

54. Tibshirani, R. Regression shrinkage and selection via the lasso. Journal of the Royal Statistical Society: Series B (Methodological) 58, 267–288 (1996).

55. Zou, H. The adaptive lasso and its oracle properties. Journal of the American statistical association 101, 1418–1429 (2006).

56. Fan, J., Gasser, T., Gijbels, I., Brockmann, M. & Engel, J. Local polynomial regression: optimal kernels and asymptotic minimax efficiency. Annals of the Institute of Statistical Mathematics 49, 79–99 (1997).

57. Beck, A. & Teboulle, M. A fast iterative shrinkage-thresholding algorithm for linear inverse problems. SIAM Journal on Imaging Sciences 2, 183–202 (2009).

58. Hyndman, R. J. & Athanasopoulos, G. Forecasting: Principles and practice (OTexts, 2018).

59. World Health Organization (WHO). https://www.who.int/docs/default-source/coronaviruse/situation-reports/20200127-sitrep-7-2019--ncov.pdf. 2020. (2020).

60. National Health and Family Planning Commission of China. Report on China’s migrant population development (China Population Publishing House, Beijing, 2017).

